# Household Transmission of Human Metapneumovirus and Human Parainfluenza Viruses 1-4 in the United States, 2022-2024

**DOI:** 10.64898/2026.08.02.26359491

**Authors:** Pavitra Roychoudhury, Anna Elias-Warren, Erica Wetzler, Hyeong Geon Kim, Kevin Kong, Hong Xie, Cassey Spring, Margaret G. Mills, Alex Harteloo, Collrane Frivold, Madison Hollcroft, Mark Drummond, Tara Hatchie, Erica Clark, Brenna Ehmen, Peter D. Han, Luis Gamboa, Sally Grindstaff, Jeremy Stone, Katherine L. Hoffman, Alexander L. Greninger, Lea M. Starita, Christina Lockwood, Janet A. Englund, Ana A. Weil, Sacha L. Reich, Richard A. Mularski, Mark A Schmidt, Jennifer L. Kuntz, Allison L. Naleway, Helen Y. Chu

## Abstract

Human metapneumovirus (HMPV) and human parainfluenza viruses (HPIV-1–4) are common causes of seasonal respiratory illness, but their household transmission dynamics remain poorly defined. We analyzed weekly symptom data and nasal swabs from a prospective household surveillance study in Washington and Oregon from June 2022–March 2024. Swabs were tested for HMPV and HPIV, and a subset underwent whole-genome sequencing. Distinct index cases occurred in 236 (23%) and 341 (36%) of 1,040 households for HMPV and HPIV, respectively. Household secondary attack rates (SAR) and median generation times were similar for HMPV (8.7%, 7 days) and HPIV (7.3%, 7 days). Sequenced samples reflected contemporaneous circulating strains during the same period, with household sequences clustering closely together. High-quality viral whole genome sequences were recovered for two or more individuals within a household in a total of 26 households for HMPV and 45 households for HPIV. Intra-household pairwise nucleotide (nt) distance ranged from 0 to 14 nt (median 0) for HMPV and 0-8 nt (median 1) for HPIV when cases occurred 0-15 days apart. Transmission occurred primarily from children to adults, emphasizing the importance of childcare-and school-associated spread and supporting child-focused prevention strategies to reduce household transmission.

## Main text

Human metapneumovirus (HMPV) and human parainfluenza viruses (HPIV) 1-4 are seasonal respiratory viruses [1,2]. They cause significant morbidity and mortality, particularly in young infants, children, older adults, immunocompromised individuals, and those with comorbid conditions [3,4]. The community burden of these viruses is less well-described in older children and working-age adults, though these viruses are known to primarily cause upper respiratory symptoms including cough, rhinitis, and lower tract symptoms including wheezing, croup, bronchiolitis and pneumonia. HMPV and HPIV are associated with school and work absenteeism in otherwise healthy populations. Reinfections throughout life are common, though infections are less severe in older children and younger adults [5].

Household studies provide an opportunity to understand the epidemiology and risk factors associated with viral transmission and the potential effect of vaccines and therapeutics on prevention of viral spread. Family studies in Tecumseh, Michigan [6] and Seattle, Washington [7] from the 1950s and 1960s showed a significant burden of these viruses in families; however, many sociologic factors have changed since this time which may have altered viral transmission dynamics. Few recent studies have described HMPV or HPIV transmission in households using molecular detection techniques or incorporating genomic sequencing to confirm chains of transmission [8,9]; more recent small-scale epidemiologic studies from the US and Japan identified school-age children as index cases, leading to onward transmission to younger children and adult family members [10].

Vaccines and monoclonal antibodies against HMPV and HPIV are in clinical development and may protect vulnerable populations from severe infection [11,12]. Baseline epidemiologic and virologic data from community-based studies provide important data on target populations for vaccination and an opportunity to understand how immunization may protect individuals from disease and onward transmission.

Here we use data from more than 1,000 households enrolled in a prospective surveillance study combined with molecular testing and viral genomic sequencing to characterize the epidemiology and household transmission dynamics of HMPV and HPIV-1–4. We estimate secondary attack rates, identify individual-and household-level risk factors for transmission, and integrate genomic data to corroborate chains of transmission. This work provides updated community-based evidence to inform prevention strategies, including vaccines and monoclonal antibody products that are in development targeting these viruses.

## Methods

### Study Design and Population

Study data were from CASCADIA, a community-based prospective cohort study of children and adults in metropolitan Seattle, Washington and Portland, Oregon, USA [13]. From June 2022 to March 2024, data were collected using remote active surveillance of respiratory viruses among enrolled households, with at least weekly at-home collection of nasal swab specimens from participants with and without symptoms. Enrollment was open to people aged ≥6 months to 49 years living in the University of Washington (UW) catchment area (King, Pierce, and Snohomish counties) or the Kaiser Permanente catchment area (northern Oregon and southern Washington). Not all household members were required to enroll in the study. The study protocol was reviewed and approved by the Kaiser Permanente Inter-regional Institutional Review Board, with reliance from University of Washington and Seattle Children’s Research Institute (45 C.F.R. part 46.114; 21 C.F.R. part 56.114).

### Data collection

Individuals were eligible for the CASCADIA study if they were between 6 months and 49 years of age at the time of enrollment. Study activities, including consent, enrollment, surveys, and nasal swab collection, were conducted at home with shipment of the samples to the University of Washington laboratories for testing. After consenting, participants were asked to complete an enrollment survey and blood draw. Participants then completed weekly surveys and collected nasal swabs regardless of symptoms, and all swabs were tested for RSV, Influenza A/B virus, and SARS-CoV-2. Nasal swabs from individuals who reported any new symptom (fever, chills, cough, shortness of breath, fatigue, sore throat, congestion or runny nose, nausea, vomiting, diarrhea, muscle or body aches, headache, and change in smell or taste, persistent pain/pressure in chest, pale/gray/blue lips, skin, or nail beds, and decreased activity and irritability/crankiness for young children) within 72 hours from their weekly swab underwent testing on a multiplex PCR panel for 26 pathogens including HMPV and HPIV as described below [13,14].

### Laboratory testing and genomic sequencing

RNA was extracted from selected nasal swab specimens using the MagNA Pure 96 DNA and viral nucleic acid small volume kit (Roche Diagnostics), with 200μL input and 50μL elution. Multiplex PCR testing was performed using the OpenArray platform (ThermoFisher) with a custom panel containing 26 targets as described previously [13,14]. This included one HMPV target (Vi99990004_po) and two HPIV targets (Vi06439642_s1 and Vi06439672_s1 detecting HPIV-1 and HPIV-2 respectively in one reaction, and Vi06439670_s1 and Vi99990005_po for HPIV-3 and HPIV-4 respectively). Where C_rt_ values are reported, the mean value for each sample was used from two replicates performed on the OpenArray assay.

Specimens positive for the HMPV and HPIV targets with C_rt_ value < 20 were selected for sequencing. For HPIV samples that were not sequenced, typing was performed by RT-PCR (primer and probe sequences in Table S1, [15]) using the AgPath-ID One Step RT-PCR kit (ThermoFisher) and PCR testing was run on the Applied Biosystems 7500 system.

Viral whole genome sequencing was performed using oligonucleotide probe capture-based enrichment for multiple respiratory viruses. Briefly, extracted RNA was converted to double-stranded cDNA, purified by bead cleanup, enzymatically fragmented, end-repaired, indexed, amplified, and purified again using the QIAseq FX DNA Library Kit (Qiagen). Hybridization capture was performed using the QIAseq xHYB Viral Respiratory Panel (Qiagen) after pooling libraries by sample C_rt_ values, with up to 6 samples in each pool. After overnight hybridization with biotinylated probes and subsequent washing to remove unbound fragments, enriched libraries were amplified and then purified by bead clean-up. Library fragment sizes were estimated by TapeStation 4200 D1000 (Agilent) and concentrations were measured by Qubit 4 Fluorometer (Invitrogen). Libraries passing quality control were sequenced on Illumina Novaseq 6000 or Nextseq 2000 instruments using a 2×150 read format. Consensus genomes were generated by using a custom bioinformatic pipeline (https://github.com/greninger-lab/revica) described previously [14]. This pipeline performs trimming of raw reads for quality, reference selection, and iterative mapping to generate a consensus genome. Assembled genomes were annotated using Viral Annotation DefineR (VADR) [16] and deposited to Genbank (Accessions available in Supplementary Table 1).

## Statistical analysis

Analysis was restricted to individuals with symptomatic HMPV and HPIV1-4 infections. We summarized demographic, clinical, and behavioral characteristics of households, index cases, and household contacts. Only multi-person households (≥2 household members enrolled) with a distinct index case were included, meaning each household contact provided one observation in the data.

An **individual illness episode** was defined as the period within which an individual’s specimen(s) were PCR-positive for HMPV or HPIV1-4 with ≤14 days separating any 2 positive specimens for the same virus or subtype of HPIV1-4. A **household illness episode** was defined as a period within which ≥1 individual illness episode(s) for the same virus or subtype of HPIV1-4 occurred in member(s) of the same household with ≤14 days separating positive specimens in the household. Single-person households were excluded from the analysis. Within households with any HMPV or HPIV infection, **index cases** were defined as the first household member with symptomatic HMPV or HPIV1-4 detected. With the exception of the cycle threshold analysis, HPIV1-2 and HPIV3-4 were grouped together in the analysis. **Co-primary index cases** were defined as two or more household members with the same date of HMPV or HPIV detection. A **household contact** was defined as a participant in the same household as an index case with a specimen(s) collected 1–14 days after the index case. **Potential secondary transmission** was defined as additional household member(s) with HMPV or HPIV detected 1 to 14 days after the index case(s) (ref for study protocol). Possible repeat infections were characterized as those infections within the same individual with two detections of the same virus separated by >14 days with no positive results in the interim period. The household **secondary attack rate** (SAR) was defined as the probability that an infection occurs among susceptible people within the same household [17]. The **detection interval** was defined as the number of days between the index case and secondary case’s first positive swab collection. See Table S1 for terms and definitions.

The primary outcome was secondary HMPV or HPIV infection among household contacts within ≤14 days of an index case. The primary model predicting the household secondary attack rate and evaluating risk factors for secondary infection included household contacts of a distinct index case. We assessed the household secondary attack rate and risk factors associated with household transmission based on log-linear Poisson regression models fitted using generalized estimating equations (GEE). Variables included in the adjusted multivariable model were determined *a priori:* household size, housing type, daycare attendance, contact age, study region, any comorbidities, and variables specific to the index case, including median C_rt_ level (for HMPV only), age, comorbidities, infection prevention measures, and care seeking. For each estimate, we obtained 95% confidence intervals and p-values using robust standard error estimates to account for clustering by household and violations of the Poisson distributional assumption.

Sensitivity analyses examined the risk of household transmission and assessed risk factors including all households with co-primary index cases and investigated the risk of HMPV and HPIV transmission restricting to households with all members enrolled (i.e. complete ascertainment). The household income cutoff of $100,000 was determined by median household income in the Seattle and Portland metropolitan areas [18,19]. The median household income in 2022 was $134,600 for Seattle and $106,000 for Portland. Based on city-specific statistics and the income survey question structure, the decision was made to make a binary cutoff of $100,000 for models included in this analysis. All analyses were conducted in R version 4.3.2.

### Sequence analysis

Study sequences were filtered to include high-quality genomes (<10% Ns that passed annotation by VADR). All publicly available genomes in NCBI Genbank as of 2025-11-19 were downloaded for HMPV (using taxonomic ID 162145 and filtered to include sequences between 12000 and 14000 nt) and HPIV (using taxonomic IDs 12730, 11216, 2560525, 2560526, 11224 and 11226 and filtered to include sequences between 13000 and 20000 nt). Trees were constructed using the Nextstrain platform [20]. Consensus sequences were aligned using augur after filtering and subsampling to include all study sequences and a random subset (5 per country-year-month) of contextual sequences downloaded from Genbank. Visualization was performed using auspice and exported for annotation in ggtree [21]. Pairwise comparison of sequences within the household was performed by aligning sequences using MAFFT [22], masking sites with ambiguities, and counting the number of pairwise nucleotide differences across the genome using the ape package [23] in R version 4.4.2.

## Results

Overall, 1,040 multi-person households were included in this analysis; 148 individuals in single-person households were excluded (Figure 1). The median reported household size was 3 (range 2-8), and 48.8% had all household members enrolled. Most households reported an income > $100,000. The age distribution of household members was as follows: 106 (3.4%) aged 6 months to one year, 256 (8.2%) 2-4 years, 943 (30.1%) aged 5-12 years, 302 (9.7%) 13-17 years, and 1521 (48.6%) 18-50 years. There were no significant differences in C_rt_ values across age groups, number of reported ARI symptoms, or by occurrence of secondary transmission (Figure S1).

**Figure 1.**
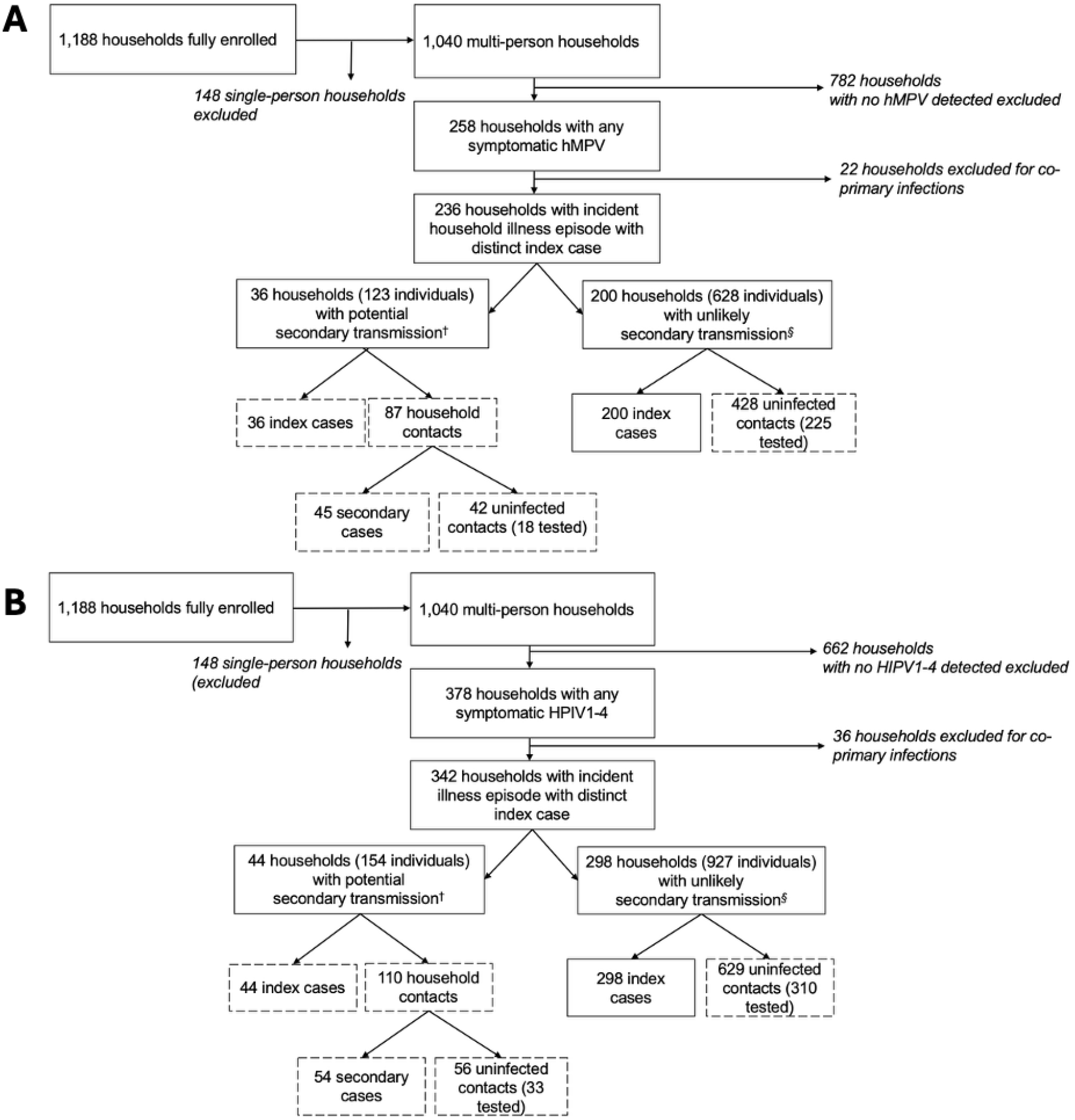
Study flow diagram for HMPV (A) and HPIV 1-4 (B), June 2022 – March 2024. See Table 1 for definition of potential vs. unlikely secondary transmission.

**Table 1.** Demographic characteristics of households with and without a secondary HMPV or HPIV infection within 14 days of a distinct index case during the incident household illness episode, Washington and Oregon, United States, 2022-2024.

|  |  | HMPV |  |  | HPIV 1-4 |  |  |
| --- | --- | --- | --- | --- | --- | --- | --- |
|  |  | Potential<br>secondary<br>transmission<sup>1</sup> | Unlikely<br>secondary<br>transmission<sup>2</sup> | Total | Potential<br>secondary<br>transmission<sup>1</sup> | Unlikely<br>secondary<br>transmission<sup>2</sup> | Total |
| *Characteristic of household* |  | (n=36) | (n=200) | (n=236) | (n=44) | (n=298) | (n=342) |
| Household density<sup>3</sup> | 2-4 individuals | 27 (75.0%) | 167 (83.5%) | 194 (82.2%) | 33 (75.0%) | 238 (79.9%) | 271 (79.2%) |
|  | 5+ individuals | 9 (25.0%) | 33 (16.5%) | 42 (17.8%) | 11 (25.0%) | 60 (20.1%) | 71 (20.8%) |
| Housing type | House/condo/townhouse | 33 (91.7%) | 193 (96.5%) | 226 (95.8%) | 43 (97.7%) | 278 (93.3%) | 321 (93.9%) |
|  | Apartment | 2 (5.6%) | 6 (3.0%) | 8 (3.4%) | 1 (2.3%) | 19 (6.4%) | 20 (5.8%) |
|  | Other | 1 (2.8%) | 0 | 1 (0.4%) | 0 | 0 | 0 |
| Household enrollment | Median<sup>4</sup> [min, max] | 3 [2, 6] | 3 [2, 6] | 3 [2, 6] | 3.5 [2, 6] | 3 [2, 6] | 3 [2, 6] |
|  | No child <5 | 22 (61.1%) | 131 (65.5%) | 153 (64.8%) | 20 (45.5%) | 173 (58.1%) | 193 (56.4%) |
|  | Child <5, but no childcare | 3 (8.3%) | 11 (5.5%) | 14 (5.9%) | 5 (11.4%) | 25 (8.4%) | 30 (8.8%) |
|  | Child <5 in childcare<sup>5</sup> | 11 (30.6%) | 58 (29.0%) | 69 (29.2%) | 19 (43.2%) | 100 (33.6%) | 119 (34.8%) |
| Income \$100,000+ | | 26 (72.2%) | 149 (74.5%) | 175 (74.2%) | 35 (79.5%) | 223 (74.8%) | 258 (75.3%) |
| Smoker in household |  | 7 (19.4%) | 24 (12.0%) | 31 (13.1%) | 7 (15.9%) | 32 (10.7%) | 39 (11.4%) |
| Study site<sup>6</sup> | KPNW | 15 (41.7%) | 87 (43.5%) | 102 (43.2%) | 17 (38.6%) | 141 (47.3%) | 158 (46.2%) |
|  | UW | 21 (58.3%) | 113 (56.5%) | 134 (56.8%) | 27 (61.4%) | 157 (52.7%) | 184 (53.8%) |
| *Characteristic of index case* |  | (n=36) | (n=200) | (n=236) | (n=44) | (n=298) | (n=342) |

<sup>1</sup> Potential secondary transmission defined as occurring when HMPV/HPIV is detected in a secondary household member 1-14 days after an index case.
<sup>2</sup> Unlikely secondary transmission defined as occurring when there is only 1 HMPV/HPIV case detected in the household OR when there are 2+ HMPV/HPIV cases in a household but secondary case detected >14 days after index case.
<sup>3</sup> Household density represents the number of household members regardless of enrollment in the study (thus, this may be greater than the number of individuals enrolled in the study if not all household members are enrolled).
<sup>4</sup> Number of participants from the household enrolled in the study
<sup>5</sup> Daycare or school attendance among at least one child <5 years in the household as reported at enrollment

|  |  |  |  |  |  |  |  |
| --- | --- | --- | --- | --- | --- | --- | --- |
| Age | Median [min, max] years | 9 [0, 49] | 9 [0, 49] | 9 [0, 49] | 6.5 [1, 39] | 8 [0, 50] | 8 [0, 50] |
|  | 6 months - 1 year | 1 (2.8%) | 3 (1.5%) | 4 (1.7%) | 3 (6.8%) | 26 (8.7%) | 29 (8.5%) |
|  | 2 - 4 years | 7 (19.4%) | 41 (20.5%) | 48 (20.3%) | 12 (27.3%) | 58 (19.5%) | 70 (20.5%) |
|  | 5 - 12 years | 17 (47.2%) | 86 (43.0%) | 103 (43.6%) | 21 (47.7%) | 118 (39.6%) | 139 (40.6%) |
|  | 13 - 50 years | 11 (30.6%) | 70 (35.0%) | 81 (34.3%) | 8 (18.1%) | 96 (32.3%) | 104 (30.4%) |
| Gender | Female | 16 (44.4%) | 115 (57.5%) | 131 (55.5%) | 16 (36.4%) | 160 (53.7%) | 176 (51.5%) |
|  | Male | 19 (52.8%) | 78 (39.0%) | 97 (41.1%) | 25 (56.8%) | 133 (44.6%) | 158 (46.2%) |
|  | Other | 1 (2.8%) | 7 (3.5%) | 8 (3.4%) | 3 (6.8%) | 5 (1.7%) | 8 (2.3%) |
| Race | Amer Indian/Alaska Native | 0 (0.0%) | 1 (0.5%) | 1 (0.4%) | 0 | 3 (1.0%) | 3 (0.9%) |
|  | Asian | 3 (8.3%) | 11 (5.5%) | 14 (5.9%) | 5 (11.4%) | 14 (4.7%) | 19 (5.6%) |
|  | Black/African American | 0 (0.0%) | 1 (0.5%) | 1 (0.4%) | 1 (2.3%) | 1 (0.3%) | 2 (0.6%) |
|  | White | 26 (72.2%) | 148 (74.0%) | 174 (73.7%) | 30 (68.2%) | 221 (74.2%) | 251 (73.4%) |
|  | Multiracial | 6 (16.7%) | 35 (17.5%) | 41 (17.4%) | 8 (18.2%) | 46 (15.4%) | 54 (15.8%) |
|  | Other | 1 (2.8%) | 4 (2.0%) | 5 (2.0%) | 0 | 13 (4.4%) | 13 (3.8%) |
|  | Hispanic | 2 (5.6%) | 20 (10.0%) | 22 (9.3%) | 5 (11.4%) | 21 (7.0%) | 26 (7.6%) |
| Any smoking |  | 2 (5.6%) | 7 (3.5%) | 9 (3.8%) | 0 | 4 (1.3%) | 4 (1.2%) |
| Any comorbidities<sup>7</sup> |  | 10 (27.8%) | 56 (28.0%) | 66 (28.0%) | 7 (15.9%) | 71 (23.8%) | 78 (22.8%) |
| Masking in public | Any | 29 (80.5%) | 148 (75.1%) | 177 (76.0%) | 28 (63.6%) | 209 (70.1%) | 237 (69.3%) |
|  | Never | 7 (19.4%) | 49 (24.9%) | 56 (24.0%) | 16 (36.4%) | 89 (29.9%) | 105 (30.7%) |
| Relative cycle threshold (C<sub>rt</sub>) | HPIV 1-2 Median [Min, Max] | 17.7 [9.4, 26.5] | 18.8 [6.7, 27.8] | 18.7 [6.7, 27.8] | 16.7 [8.4, 26.2] | 20.7 [2.3, 31.2] | 20.3 [2.3, 31.2] |
|  | HPIV 3-4 Median [Min, Max] | -- | -- | -- | 15.1 [7.3, 25.7] | 20.3 [8.7, 35.7] | 19.7 [7.3, 35.7] |
|  | ≤ median<sup>8</sup> | 23 (63.9%) | 97 (48.5%) | 120 (50.8%) | 13/17 (76.1%) | 63/141 (44.7%) | 76/158 (48.1%) |
|  | ≤ median | -- | -- | -- | 17/27 (63.0%) | 82/172 (47.7%) | 99/199 (49.7%) |
| Duration of viral detection ≥ 1 week |  | 0 (0.0%) | 0 (0.0%) | 0 (0.0%) | 1 (2.3%) | 2 (0.7%) | 3 (0.9%) |
| Presence of viral codetection | Any | 10 (27.8%) | 64 (32.0%) | 74 (31.4%) | 14 (31.6%) | 79 (26.5%) | 93 (27.2%) |
|  | SARS-CoV-2 | 2 (5.6%) | 3 (1.5%) | 5 (2.1%) | 4 (9.1%) | 5 (1.7%) | 9 (2.6%) |
|  | Influenza | 0 (0.0%) | 1 (0.5%) | 1 (0.4%) | 0 | 2 (0.7%) | 2 (0.6%) |

<sup>7</sup> Comorbidities include asthma, chronic obstructive pulmonary disease (including chronic bronchitis and emphysema), sleep apnea, heart disease, congenital heart disease, heart failure, down syndrome, hypertension, diabetes, liver condition, weak or failing kidneys, cancer or malignancy, arthritis, stroke, deep vein thrombosis or pulmonary embolism, sickle cell disease or thalassemia, weakened immune system, depression, anxiety, thyroid issues, or other health diagnosis.
<sup>8</sup> HMPV: 18.6 C<sub>rt</sub>; HPIV 1-2: 20.3 C<sub>rt</sub>; HPIV 3-4: 19.7 C<sub>rt</sub>

|  |  |  |  |  |  |  |  |
| --- | --- | --- | --- | --- | --- | --- | --- |
|  | Rhinovirus | 4 (11.1%) | 30 (15.0%) | 34 (14.4%) | 7 (15.9%) | 37 (12.4%) | 44 (12.9%) |
|  | Adenovirus | 3 (8.3%) | 16 (8.0%) | 19 (8.1%) | 0 | 23 (7.7%) | 23 (6.7%) |
|  | RSV | 0 | 4 (2.0%) | 4 (1.7%) | 2 (4.5%) | 3 (1.0%) | 5 (1.5%) |
|  | Enterovirus | 0 (0.0%) | 6 (3.0%) | 6 (2.5%) | 0 | 5 (1.7%) | 5 (1.5%) |
|  | Human coronavirus | 1 (2.8%) | 12 (6.0%) | 13 (5.5%) | 1 (2.3%) | 11 (3.7%) | 12 (3.5%) |
|  | HMPV | NA | NA | NA | 0 | 5 (1.7%) | 5 (1.5%) |
|  | HPIV | 0 (0.0%) | 5 (2.5%) | 5 (2.1%) | NA | NA | NA |
| Any ARI symptom(s)<sup>9</sup> | 1+ ARI symptoms | 36 (100%) | 200 (100%) | 236 (100%) | 44 (100%) | 297 (100%) | 341 (100%) |
|  | 2+ ARI symptoms | 29 (80.6%) | 154 (77.0%) | 183 (77.5%) | 40 (90.9%) | 236 (79.2%) | 276 (80.7%) |
|  | Cough and/or rhinorrhea | 35 (97.2%) | 196 (98.0%) | 231 (97.9%) | 42 (95.5%) | 285 (96.0%) | 327 (95.9%) |
| Any care seeking during illness<sup>10</sup> |  | 5 (13.9%) | 27 (13.5%) | 32 (13.6%) | 6 (13.6%) | 36 (12.1%) | 42 (12.3%) |
| Any behavior change to reduce transmission in the air |  | 15 (41.7%) | 61 (30.5%) | 76 (32.2%) | 14 (31.8%) | 72 (24.2%) | 86 (25.1%) |
| Any behavior change to reduce transmission on surfaces |  | 15 (41.7%) | 55 (27.5%) | 70 (29.7%) | 13 (29.5%) | 72 (24.2%) | 85 (24.9%) |
| *Characteristics of household contacts* |  | (n=87) | (n=428) | (n=515) | (n=110) | (n=629) | (n=739) |
| Age | Median [min, max] years | 37 [0, 49] | 37.5 [0, 49] | 37 [0, 49] | 35.5 [0, 49] | 36 [0, 49] | 36 [0, 49] |
|  | 6 months - 1 year | 2 (2.3%) | 6 (1.4%) | 8 (1.6%) | 4 (3.6%) | 14 (2.2%) | 18 (2.4%) |
|  | 2 - 4 years | 4 (4.6%) | 21 (4.9%) | 25 (4.9%) | 8 (7.3%) | 39 (6.2%) | 47 (6.4%) |
|  | 5 - 12 years | 28 (32.2%) | 113 (26.4%) | 141 (27.4%) | 22 (20.0%) | 159 (25.3%) | 181 (24.5%) |
|  | 13 - 50 years | 53 (60.9%) | 288 (67.3%) | 341 (66.2%) | 76 (69.0%) | 417 (66.3%) | 493 (66.7%) |
| Gender | Female | 54 (62.1%) | 232 (54.2%) | 286 (55.5%) | 59 (53.6%) | 364 (57.9%) | 423 (57.2%) |
|  | Male | 31 (35.6%) | 185 (43.2%) | 216 (41.9%) | 48 (43.6%) | 248 (39.4%) | 296 (40.1%) |
|  | Other | 2 (2.3%) | 11 (2.6%) | 13 (2.6%) | 3 (2.7%) | 17 (2.7%) | 20 (2.8%) |
| Race | American Indian/Alaska | 0 (0.0%) | 0 (0.0%) | 0 (0.0%) | 0 | 3 (0.5%) | 3 (0.4%) |
|  | Asian | 6 (6.9%) | 28 (6.5%) | 34 (6.6%) | 18 (16.4%) | 38 (6.0%) | 56 (7.6%) |
|  | Black/African American | 0 (0.0%) | 1 (0.2%) | 1 (0.2%) | 0 | 7 (1.1%) | 7 (0.9%) |
|  | Native Hawaiian/Pacific | 1 (1.1%) | 1 (0.2%) | 2 (0.4%) | 0 | 1 (0.2%) | 1 (0.1%) |

<sup>9</sup> ARI, acute respiratory illness symptom reported within $\pm 7$ days of the individual HMPV illness episode's first positive specimen collection, including fever, cough, sore throat, shortness of breath, myalgia, and/or rhinorrhea.
<sup>10</sup> Seeking health care from a healthcare provider during illness, self-reported
<sup>11</sup> Includes masking, sleeping separately, covering cough/sneeze
<sup>12</sup> Includes handwashing, cleaning, disinfecting

|  |  |  |  |  |  |  |  |
| --- | --- | --- | --- | --- | --- | --- | --- |
|  | White | 66 (75.9%) | 348 (81.3%) | 414 (80.4%) | 85 (77.3%) | 507 (80.6%) | 592 (80.1%) |
|  | Multiracial | 12 (13.8%) | 42 (9.8%) | 54 (10.5%) | 6 (5.5%) | 55 (8.7%) | 61 (8.3%) |
|  | Other | 2 (2.3%) | 8 (1.9%) | 10 (1.9%) | 1 (0.9%) | 18 (2.8%) | 19 (2.6%) |
|  | Hispanic | 7 (8.0%) | 36 (8.4%) | 43 (8.3%) | 8 (7.3%) | 40 (6.4%) | 48 (6.5%) |
| Any smoking |  | 5 (5.7%) | 21 (4.9%) | 26 (5.0%) | 7 (6.4%) | 31 (4.9%) | 38 (5.1%) |
| Any comorbidities<sup>7</sup> |  | 35 (40.2%) | 181 (42.3%) | 216 (41.9%) | 52 (47.3%) | 275 (43.7%) | 327 (44.2%) |
| Masking in public | Any | 72 (82.8%) | 340 (79.4%) | 412 (80.0%) | 85 (77.3%) | 494 (78.5%) | 579 (78.3%) |
|  | Never | 15 (17.2%) | 88 (20.6%) | 103 (20.0%) | 25 (22.7%) | 135 (21.5%) | 160 (21.7%) |
| Uninfected contacts | N | 42 | 428 | 470 | 56 | 629 | 739 |
|  | Tested | 24 (57.1%) | 225 (52.6%) | 249 (53.0%) | 33 (58.9%) | 310 (49.4%) | 343 (50.2%) |
|  | Not tested | 18 (42.9%) | 203 (47.4%) | 221 (47.0%) | 23 (41.1%) | 319 (50.9%) | 340 (49.8%) |

**Table 2.** Factors associated with household transmission of HMPV within ≤14 days of a distinct index case, Washington and Oregon, United States, 2022-2024 (n=515 contacts). RR = relative.

|  |  | % of contacts infected<sup>15</sup> | Unadjusted RR (95% CI) | Adjusted RR (95% CI) | p-value<sup>16</sup> |
| --- | --- | --- | --- | --- | --- |
| *Characteristic of household* |  |  |  |  |  |
| Household density<sup>17</sup> | 2-4 individuals | 8.2% (32/389) | - | - | 0.727 |
|  | 5+ individuals | 10.3% (13/126) | 1.25 (0.56, 2.79) | 1.16 (0.50, 2.66) |  |
| Housing type | House, condo, or | 8.4% (42/501) | - | - | **0.009** |
|  | Other<sup>18</sup> | 21.4% (3/14) | 2.56 (1.06, 6.17) | 3.31 (1.34, 8.18) |  |
| Childcare attended by child $<5$ years in household<sup>19</sup> | No child $<5$ | 8.3% (26/315) | - | - | |
| | Child $<5$, but no | 13.9% (5/36) | 1.68 (0.57, 4.96) | 1.15 (0.31, 4.31) | 0.799 |
| | Child $<5$ in childcare | 8.5% (14/164) | 1.03 (0.50, 2.15) | 0.82 (0.30, 2.28) | |
| Study site<sup>20</sup> | KPNW | 8.9% (19/213) | - | - | 0.801 |
|  | UW | 8.6% (26/302) | 0.97 (0.48, 1.93) | 1.09 (0.57, 2.07) |  |
| *Characteristic of household contact* |  |  |  |  |  |
| Age | 6 mo - 4 years | 20.7% (6/29) | 2.69 (1.23, 5.90) | 3.70 (1.23, 11.08) |  |
|  | 5 - 12 years | 8.9% (12/135) | 1.16 (0.63, 2.11) | 1.40 (0.67, 2.92) |  |
|  | 13 - 50 years | 7.7% (27/351) | - | - | 0.064 |
| Any comorbidities<sup>21</sup> | Yes | 8.8% (19/216) | 1.01 (0.60, 1.72) | 1.45 (0.75, 2.80) | 0.272 |
|  | No | 8.7% (26/299) | - | - |  |
| *Characteristic of index case* |  |  |  |  |  |
| Age | 6 mo - 4 years | 8.0% (10/125) | 1.03 (0.43, 2.51) | 1.50 (0.44, 5.05) |  |
|  | 5 - 12 years | 9.8% (23/235) | 1.26 (0.59, 2.70) | 1.60 (0.76, 3.39) | 0.461 |
|  | 13 - 50 years | 7.7% (12/155) | - | - |  |
| Relative cycle threshold ($C_{rt}$) ≤ median (18.7 $C_{rt}$) | Yes | 11.9% (28/236) | 1.95 (0.99, 3.82) | 2.06 (1.09, 3.91) | **0.027** |
|  | No | 6.1% (17/279) | - | - |  |
| Presence of viral codetection | Yes | 6.7% (11/163) | 0.70 (0.34, 1.44) | 0.69 (0.33, 1.43) | 0.320 |
|  | No | 9.7% (34/352) | - | - |  |
| Any care seeking during illness<sup>24</sup> | Yes | 15.4% (10/65) | 1.98 (0.76, 5.13) | 1.40 (0.58, 3.40) | 0.451 |
|  | No | 7.8% (35/450) | - | - |  |
| Any behavior to reduce transmission in the air<sup>25</sup> | Yes | 13.5% (21/156) | 2.01 (1.03, 3.93) | 1.28 (0.63, 2.59) | 0.451 |
|  | No | 6.7% (24/359) | - | - |  |
| Any behavior to reduce transmission on surfaces<sup>26</sup> | Yes | 14.4% (21/146) | 2.21 (1.14, 4.30) | 1.76 (0.88, 3.50) | 0.110 |
|  | No | 6.5% (24/369) | - | - |  |

**Table 3.** Factors associated with household transmission of HPIV within ≤14 days of a distinct index case, Washington and Oregon, United States, 2022-2024 (n=739 contacts)

|  |  | % of contacts infected<sup>15</sup> | Unadjusted relative risk (95% CI) | Adjusted relative risk (95% CI) | p-value<sup>16</sup> |
| --- | --- | --- | --- | --- | --- |
| *Characteristic of household* |  |  |  |  |  |
| Household density<sup>17</sup> | 2-4 individuals | 6.8% (37/545) | - | - | 0.710 |
|  | 5+ individuals | 8.8% (17/194) | 1.29 (0.67, 2.50) | 1.14 (0.58, 2.24) |  |
| Housing type | House, condo, or | 7.5% (53/703) |  | - | 0.346 |
|  | Other<sup>18</sup> | 2.7% (1/36) | 0.37 (0.05, 2.67) | 0.37 (0.05, 2.95) |  |
| Childcare attended by child <5 years in household<sup>19</sup> | No child <5 | 6.0% (23/383) | - | - | 0.983 |
|  | Child <5, but no | 7.7% (6/78) | 1.28 (0.50, 3.29) | 1.08 (0.36, 3.23) |  |
|  | Child <5 in childcare | 9.0% (25/278) | 1.50 (0.81, 2.66) | 0.98 (0.42, 2.30) |  |
| Study site<sup>20</sup> | KPNW | 5.8% (20/342) | - | - | 0.184 |
|  | UW | 8.6% (34/397) | 1.46 (0.81, 2.66) | 1.50 (0.82, 2.73) |  |
| *Characteristic of household contact* |  |  |  |  |  |
| Age | 6 mo - 4 years | 15.3% (9/59) | 2.27 (1.18, 4.37) | 3.18 (1.40, 7.24) | **0.016** |
|  | 5 - 12 years | 6.3% (11/175) | 0.93 (0.47, 1.84) | 1.03 (0.54, 1.97) |  |
|  | 13 - 50 years | 6.7% (34/505) | - | - |  |
| Any comorbidities<sup>21</sup> | Yes | 7.6% (25/327) | 1.09 (0.65, 1.81) | 1.34 (0.74, 2.42) | 0.328 |
|  | No | 7.0% (29/412) | - | - |  |
| *Characteristic of index case* |  |  |  |  |  |
| Age | 6 mo - 4 years | 8.3% (17/205) | 1.85 (0.78, 4.41) | 2.01 (0.77, 5.27) | 0.233 |
|  | 5 - 12 years | 8.4% (28/333) | 1.88 (0.84, 4.21) | 1.93 (0.86, 4.35) |  |
|  | 13 - 50 years | 4.5% (9/201) | - | - |  |
| Presence of viral codetection | Yes | 9.0% (19/211) | 1.36 (0.72, 2.55) | 1.26 (0.65, 2.47) | 0.492 |
|  | No | 6.6% (35/528) | - | - |  |
| Any care seeking during illness<sup>24</sup> | Yes | 8.6% (7/81) | 1.21 (0.53, 2.79) | 1.15 (0.51, 2.59) | 0.739 |
|  | No | 7.1% (47/658) | - | - |  |
| Any behavior to reduce transmission in the air<sup>25</sup> | Yes | 10.4% (20/192) | 1.68 (0.91, 3.10) | 1.48 (0.54, 4.05) | 0.449 |
|  | No | 6.2% (34/547) | - | - |  |
| Any behavior to reduce transmission on surfaces<sup>26</sup> | Yes | 9.5% (17/179) | 1.44 (0.75, 2.74) | 1.15 (0.41, 3.22) | 0.792 |
|  | No | 6.6% (37/560) | - | - |  |

### HMPV and HPIV have similar household secondary attack rates

A distinct index case of HMPV was detected in 236 (23%) of 1040 households during the two-year surveillance period; 36 (15%) households had potential secondary transmission within 14 days of a distinct index case. The household secondary attack rate (SAR) for HMPV was 8.7% (95% CI: 6.3-12.2%), and the median detection interval between index and secondary cases was 7 days (IQR: 7-7), which is driven by the swab collection cadence of ∼7 days. A distinct index case of parainfluenza was detected in 342 (36%) of 1040 households during the two-year surveillance period; 44 (13%) households had potential secondary transmission within 14 days of a distinct index case. For HPIV, the household SAR was 7.3% (95% CI: 5.5-9.8%), and the median detection interval between index and secondary cases was 7 days (IQR: 6-7). The majority of parainfluenza household secondary transmission was HPIV-3 (n=21, 46.7%), followed by HPIV-4 (n=9, 20%), HPIV-1 (n=8, 17.8%), and HPIV-2 (n=7, 15.6%). Of 54 people with secondary infections, 9 (54%) samples either could not be typed or the type was undetermined.

### Differences in demographics and risk factors for household transmission between HMPV and HPIV

For both HPIV and HMPV, the proportion of households with more than five individuals was 25% in households with secondary transmission. However, in households without secondary transmission, 17% of households with HMPV had more than 5 people compared to 20% for HPIV. There were similar proportions with children under five in childcare (31% vs. 29% and 43% vs 34%, respectively for HMPV and HPIV) among households with and without secondary transmission. For HMPV, the median age of the index case was 9 years old in households both with and without secondary transmission, and 7 vs 8 years old for HPIV, respectively. In terms of the household contacts, the median age was similar in both groups for HMPV (37 vs. 37.5 years) and HPIV (35 vs. 36 years), though, for HMPV, the proportion of females was higher among household contacts with secondary transmission compared to those without (62% vs. 54%, respectively).

On multivariate analysis of factors associated with household transmission of HMPV, index cases with C_rt_ below the median were associated with increased risk of transmission (aRR: 2.06, 95% CI: 1.09, 3.91), as was housing type classified as ‘other’ (aRR: 3.31, 95% CI: 1.34, 8.18); factors including age of index, household density, and presence of children in childcare were not associated with increased risk of transmission. We conducted two sensitivity analyses: inclusion of co-primary cases (Supplemental Table 1) and excluding households without complete case ascertainment (Supplemental Table 2). The effect of C_rt_ on risk of transmission was seen with the first sensitivity analysis and, in contrast to the primary analysis, household contacts ≤12 years old were more likely to have a secondary infection (p=0.034); no factors were associated with risk of transmission in the second sensitivity analysis.

For HPIV, there was a higher proportion of index cases aged 2-4 years old (27% vs 19%) and a lower proportion of index cases aged 13-50 years among households with secondary transmission compared to those without secondary transmission (18% vs 33%). The proportion of female index cases in households with secondary transmission was lower compared to index cases in households without secondary transmission (41% vs 55%). The median age of household contacts was approximately 34-35 years old in both groups.

In a multivariate analysis, household contacts aged 6 months-4 years had 3.18 times higher risk of secondary transmission (95% CI: 1.40, 7.24) when compared to contacts aged 13-50 years after adjusting for covariates. No other factors were significantly associated with HPIV household transmission, though we did not evaluate associations with C_rt_ because of differences in the laboratory assays. In both sensitivity analyses, household contacts 6 months to 12 years continued to have increased risk for secondary household transmission, as in the primary analysis; however, restricting to households with complete ascertainment revealed that index age 6 months to 12 years was associated with at least fourfold higher risk of household transmission compared to those 13 to 50 years (p=0.030).

### Frequent co-detection of other respiratory viruses

Viral co-detections were identified in 31.4% of HMPV and 27.2% of HPIV index case infections (Table 1). Among HMPV index cases, the most frequently co-detected pathogens were rhinovirus (14.4%), adenovirus (8.1%), and seasonal human coronaviruses (5.5%), with less frequent detection of enterovirus (2.5%), RSV (1.7%), influenza (0.4%), and SARS-CoV-2 (2.1%). Similar patterns were observed for HPIV, where rhinovirus (12.9%), adenovirus (6.7%), and seasonal coronaviruses (3.5%) were the most common co-detections. HPIV was detected in 2.1% of HMPV index cases, while HMPV was detected in 1.5% of HPIV index cases.

Co-detection patterns did not differ substantially between households with and without secondary transmission for either virus. The only notable differences, though not statistically significant, were in HPIV index cases where there was a higher frequency of SARS-CoV-2 co-detection (9.1% vs 1.7%), and a lower frequency of adenovirus co-detection (0% vs. 7.7%) in cases with secondary transmission. However, data were sparse, so associations for individual viruses were not tested in the multivariable models.

Since sequencing was performed using a hybridization capture-based method enriching for respiratory viruses (Appendix), genomes of coinfecting viruses were recovered in 46 samples along with HMPV or HPIV1-4.

### Distribution of HMPV clades and HPIV types broadly matched circulating viruses during the same period

A total of 135 high-quality HMPV consensus sequences were recovered from 135 participants from 116 households (Table S2). Study sequences represented 35.5% of all publicly available HMPV sequences (n=380) during the study period, and 68.9% of sequences from Washington and Oregon. The distribution of HMPV clades identified among study samples generally mirrored globally circulating clades during the study period (Figure 2A) with A.2.2.2/A2b2 being the most prevalent, followed by B2, A.2.2.1/A2b1, and B1 however the CASCADIA subset had a higher proportion of B2 sequences than the non-CASCADIA set (34.1% vs. 13.1%). A total of 249 high-quality HPIV consensus sequences were recovered from 238 unique participants from 207 households (Table S4). These included 73 HPIV-1, 36 HPIV-2, 65 HPIV-3, and 72 HPIV-4 (61 HPIV-4a, 11 HPIV-4b) sequences. Study sequences represented 42.4% of all publicly available sequences in Genbank during the study period and 87.7% of sequences from Washington and Oregon (Figure 2B). Although globally circulating HPIV types were detected contemporaneously in the CASCADIA cohort, the proportion of HPIV-3 was higher among non-CASCADIA sequences, while CASCADIA sequences were more evenly distributed across types with a higher share of HPIV-1 and HPIV-4 (Figure S2). The HPIV-3 sequences in the non-CASCADIA group were primarily from Russia (53.7%), Asia (18.8%), and Europe (14.2%).

**Figure 2.**
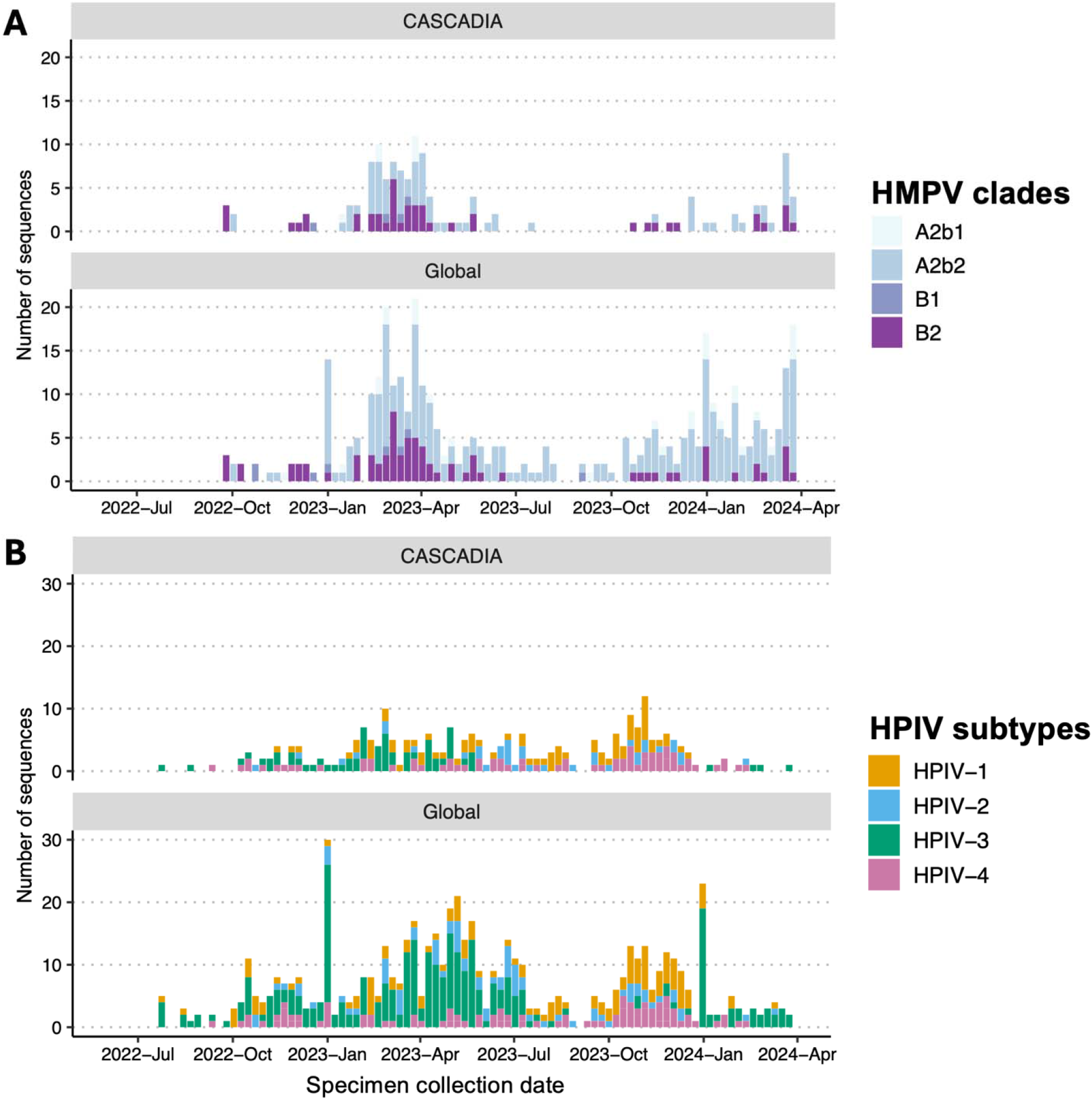
Genetic diversity of study samples compared to global sequences reflected in HMPV clades (A) and HPIV types (B) represented among study samples vs globally available sequences during the same time period.

### Genomic analysis of HMPV and HPIV transmission in households

High-quality viral whole genome sequences were recovered for two or more individuals within a household in a total of 26 households for HMPV and 45 households for HPIV.

Among the 26 households with sequenced HMPV cases, nearly all (24 out of 26) had cases occurring 0-15 days apart and among these households with HMPV, the within-household pairwise nucleotide (nt) distance between sequences ranged from 0 to 14 nt (median 0 nt). In the remaining two households, cases occurred 231 and 388 days apart and the genomic data was consistent with independent introductions.

Among the 45 households with sequenced HPIV cases, 30 households (67%) had sequenced cases ≤15 days apart. Most of these households (28 of 30) had a single HPIV type detected with near-identical genomes (0-8 pairwise differences across the genome, median 1); the remaining two households had multiple HPIV types, likely from separate introductions. In households where cases occurred more than 15 days apart, nearly all (13 out of 15) involved infection with a different HPIV type. The remaining two households had genotyped HPIV-3 cases 99 and 112 days apart with pairwise distances of 31 and 34 nucleotides, respectively, all in children (Figure 3). One household that was enrolled over multiple seasons experienced likely transmission of HPIV-3 from child to adult over 6 days supported by 100% pairwise identity of consensus genomes, and two separate HPIV-1 introductions 7 months apart.

**Figure 3.**
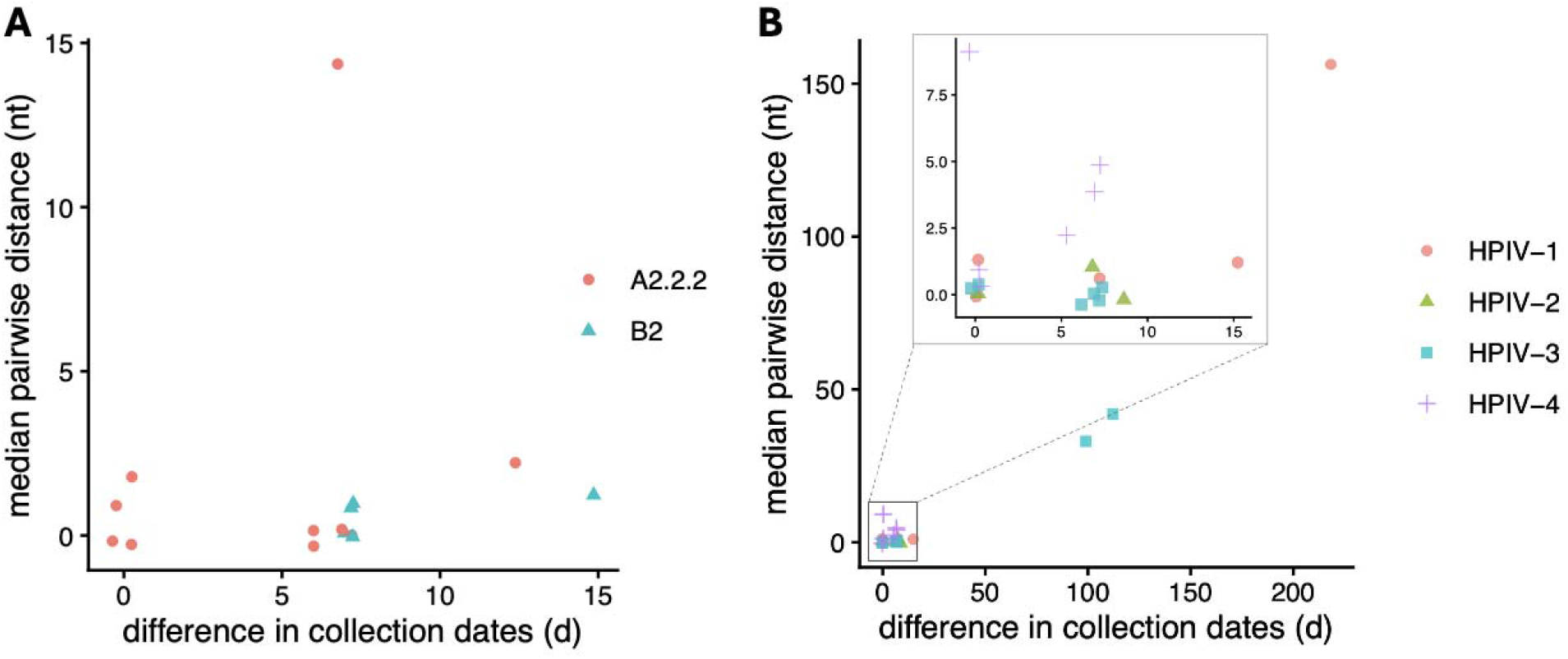
Median pairwise genetic distance between within-household case pairs. Each point represents a pair of sequenced cases of HMPV (A) or HPIV (B) within the same household, colored by HMPV clade or HPIV type. The inset in panel B shows an expanded view of pairs collected within 15 days of each other.

## Discussion

Over two years of active home-based weekly surveillance for respiratory illness in families in the United States, we observed a substantial burden of HMPV and HPIV infections. In total, one-quarter and one-third of households had an HMPV or HPIV infection, respectively, over the study period, with transmission to a secondary household contact in approximately 7% and 9% of households, respectively. The direction of transmission is typically from child to adult, suggesting the directionality of infections were contracted initially through child communal settings such as daycare or school exposures with subsequent onward infections in caregivers. This study included over 1,000 multi-person households and identified factors that are associated with increased risk of transmission, including HMPV semi-quantitative viral load. The results of genomic sequencing further validate these observations.

There are few studies of HMPV and HPIV transmission in the community setting, though HMPV has been well-described as the source of nosocomial outbreaks in adult and pediatric inpatient facilities [24,25]. A household transmission study conducted in 1,394 families with school-aged children in Wisconsin found a lower proportion of households (5%) with HMPV infection compared to this study, though similar secondary infection rates of 12% [8]. This difference may be due to the differences in study design; the case-ascertained design in Wisconsin relied on identification of a symptomatic index case for enrollment, as compared to the prospective observational study design used for this study.

A clinic-based study conducted in Japan of family members of an index HMPV case identified a serial interval based on symptoms of 4-5 days, which is consistent with the findings of this study where we identified a median detection interval based on laboratory results of 7 days, which is likely driven by our weekly swab collection cadence [9]. This is also largely concordant with a nosocomial outbreak of HMPV in a pediatric hematology-oncology ward, where the estimated incubation period was 7-9 days [26].

We identified a higher semiquantitative viral load (lower C_rt_ value) in the index case as a risk factor for secondary HMPV transmission; because of the differences in assay types used for HPIV, we could not evaluate the role of viral load in HPIV transmission. Though this has not been previously described for HMPV, household transmission studies of influenza virus, RSV, and SARS-CoV-2 have identified higher viral load of the index case as risk factors for secondary transmission. Unlike other studies in the past, we did not find that the index or secondary case age was significantly associated with transmission. The Wisconsin study found that younger age of the index and older age of the secondary case was associated with transmission; this was largely in line with our findings, where the median age of the index case was 9 years old, while the median age of the secondary case was 33 years old. In prior studies of household transmission of respiratory viruses in rural Nepal, the chain of transmission was observed to be from school-aged to younger children in the household [27]; we did not find this to be the case, likely due to differences in household density and other factors that differ between the United States and Nepal.

For HPIV household transmission, we found a similar proportion of households with secondary transmission (7%). Overall, these findings are similar to other respiratory viruses in the CASCADIA study, including RSV with 10% and EV-D68 with 13.6% households with secondary transmission [28,29].

Household contacts between 6 months and 12 years are more likely to have a secondary household infection than their older counterparts. This study contributed significantly to the number of publicly available sequences for HMPV and HPIV in Genbank, representing a large proportion of sequences globally during the study period. Genomic analysis showed that multiple introductions into the household are infrequent when cases occur within a 2-week window for both HMPV and HPIV. Viruses circulating within a household tend to be highly similar with a median of 0 pairwise nucleotide differences at the consensus genome level.

Limitations of our study include the lack of testing of asymptomatic swabs collected from individuals during their routine weekly surveillance. It is possible that asymptomatic introduction events were missed, and therefore, secondary symptomatic cases were misclassified as index cases. Second, because of the weekly swabbing cadence, we could not assess the relationship between peak viral load and risk of transmission; daily swabbing data would be important to consider in future studies to fully evaluate the relationship between viral load and risk of transmission. Additionally, not all households had complete enrollment of their members; we did conduct an additional sensitivity analysis restricting to only the households with complete case ascertainment and found the effect of viral load on transmission was no longer significant, likely due to smaller sample sizes. Finally, we were only able to sequence a limited number of the overall samples due to lower viral load, which limited our ability to completely map chains of transmission in these studies, though it is likely that illness episodes with higher viral load, where sequences are available, are associated with risk of transmission.

Overall, this study provides evidence that HMPV and HPIV cause a substantial burden of disease in households with young children in a contemporaneous study in the United States, and that transmission appears to be from young children to adults. A strategy of early post-exposure prophylaxis should be considered for other viruses that can cause infection in vulnerable household members. These studies also permit identification of high-risk priority groups to inform vaccine development, and implementation strategies so that as vaccines become available, they can be administered to individuals who may be the drivers of household transmission, in order to break chains of community transmission and have the greatest health impact. These results support the value proposition for vaccines targeted for specific populations for reduction both in burden of disease and reduction of community transmission.

## Data Availability

All data produced in the present work are contained in the manuscript.

## Acknowledgements

The authors wish to acknowledge the following individuals for supporting this study. *Kaiser Permanente Center for Health Research*: Deralyn Almaguer, David Amy, Britt Ash, Allison Bianchi, Cassandra Boisvert, Stacy Bunnell, Joseph Cerizo, Evelin Coto, Phil Crawford, Robin Daily, Lantoria Davis, Stephen Fortmann, Kendall Frimodig, Lisa Fox, Holly Groom, Tarika Holness, Matt Hornbrook, Terry Kimes, Keelee Kloer, Dorothy Kurdyla, Bryony Melcher, John Ogden, Jennifer Rivelli, Katrina Schell, Emily Schield, Meagan Shaw, Martin Simer, Britta Torgrimson-Ojerio, Alexandra Varga, Mica Werner, Neil Yetz, Rebecca Ziebell; *University of Washington*: Ariana Magedson, Denise McCulloch, Natalie Lo, Kyle Luiten, Devon McDonald, Sarah Cox, Jenni Logue, Jean Mernaugh, Melissa MacMillan, Kat Hoffman, Grace Marshall, Daniel Nguyen, Zarna Marfatia, Amanda Casto, Chidozie Iwu, Julia Bennett, Jordan Opsahl, Kathryn McCaffrey, David Reinhart, Ben Cappodano, Sarah Heidl, Zack Acker, Lani Regelbrugge, Leslie Rodriguez-Salas, Ailyn Perez, Sean Ellis, Hanna Edgar; *Seattle Children’s Hospital:* Dallas Haws, Hanna Grioni, Josh Sanders, Irem Onalan, Laura Ostrina Restrepo.

